# Male nurses’ adaptation experiences after turnover to community institutions in Korea

**DOI:** 10.1101/2023.04.24.23288853

**Authors:** Ja-Sook Kim, Suhyun Kim, Hyang-In Cho Chung

**Affiliations:** Department of Nursing, Kunsan National University, Gunsan, Jeollabuk-do, Republic of Korea; Department of Nursing, Nambu University, Gwangju, Republic of Korea; College of Nursing, Chonnam National University, Gwangju, Republic of Korea

## Abstract

We aimed to develop a substantive theory according to the associations between adaptation experience-related factors identified in male nurses after turnover to community institutions. From April through August 2019, data were collected through direct observations and in-depth interviews of 22 male nurse participants who were recruited purposively and analyzed simultaneously with the method proposed by Strauss and Corbin. Furthermore, 29 subcategories were derived from 11 categories, including: (1) leaving the clinical sector and changing jobs, (2) shaking while settling, (3) characteristics of the new job, (4) personal disposition, (5) support system, (6) finding my place, (7) solidifying my place, (8) demonstrating my professional competence, (9) stable settlement in my place, (10) preparing for a better future, and (11) still confused. The core category was identified as “putting down roots in another place for myself.” The verification of this theory in this study’s results indicates a need for research into the evaluation and development of professional development programs and related policies to provide support to male nurses who are pursuing opportunities in community institutions to maintain their nursing identity and further their efforts for developing a nursing specialty. Keywords: nurses, male, turnover, adaptation, grounded theory, qualitative research

## Introduction

Because of rapidly changing medical policies and changes in awareness about nursing expertise [1], more than 20,000 male nurses have been qualified in Korea and constitute approximately 5% of all nurses in the country [2]. The number of male students entering the nursing field will likely increase with an increase in youth unemployment rates [3]. Furthermore, the increased number of male nurses has not only changed sociocultural gender stereotypes in the nursing profession but also has expanded the availability of nursing human resources, improved working conditions, and expanded work fields [4]. The demand for male nurses is expected to increase because of these advantages. However, male nurses in the clinical sector experience difficulties because of medical–environmental factors, characteristics of nursing organizational culture, and gender characteristics [5–8] and face discrimination as a minority [5], as well as rigid organizational cultures, irrational special task assignments, and female-oriented promotion systems [6,7]. Such experiences have led to some male nurses leaving the clinical sector.

Male nurses who left the clinical sector accounted for approximately half of all male nurses in 2020 [9]. Nurses who leave the clinical sector move to various fields, such as nursing education, public health, firefighting, industry, and community health promotion [10]. The transfer of male nurses to community organizations can help improve public awareness about male nurses [11]. Furthermore, despite posing another challenge, the migration of nurses to community organizations constitutes an opportunity to expand the nursing field and enhance professionalism [12,13]. In particular, as the importance of health promotion and disease prevention is emphasized—with consideration of population aging, changes in disease patterns toward increasing chronic disease rates, and increasing interest in healthy lifestyles—the roles of community nurses are expected to gain prominence in the future [14]. Therefore, it is necessary to understand how male nurses who move to community institutions adapt to their new contexts. Moreover, it is important to help these nurses expand their professional niches within the community while maintaining their identities and expertise. Above all, this study holds significant value in cultures like Korea and Japan, where the profession of male nursing is not commonly accepted [15].

Turnover refers to the transition of workers from one workplace to another [16]. The adaptation process after changing jobs refers to the process whereby male nurses maintain harmonious relationships and a sense of stability despite situational or environmental changes [17]. In other words, understanding the adaptation process of male nurses after they move to community institutions will help increase the work adaptability and professionalism of nurses who move to new occupational fields and improve the competency of professional nurses in the community.

The experiences of nurses working as paramedics [18], correctional facility nurses [19], and elementary school health teachers [20] have been investigated. However, few studies have comprehensively analyzed conditions related to the adaptation process after transitioning to new professional realms. Existing studies, in a fragmentary manner, have focused on the experiences associated with individual occupations. Therefore, this study was conducted to comprehensively analyze male nurses’ adaptation experiences after moving to community institutions and understand the processes by which male nurses change and adapt from the clinical setting to community institutions in terms of situational contexts. Furthermore, the study investigated how various conditions interact in the transition process, and this information was used to derive a substantive theory about the adaptative experience.

The grounded theory method forms an empirical model for a process of change that has not yet been clearly identified and can be appropriately used in research to explore concepts or fields that require additional explanations, examples, or definitions [21]. Therefore, this study used the grounded theory method to ask the research question, “What are the adaptation experiences of male nurses after moving to community institutions?”

## Materials and methods

### Study design

To present a substantive theory, this qualitative study applied the grounded theory method of Corbin and Strauss [21] to identify the relationship between male nurses’ experiences and related conditions in the adjustment process after leaving a job.

### Participants

Eligible participants were male nurses younger than 40 years of age, with experience in working at medical institutions, who had moved to community institutions and had more than 6 months of work experience in the community institution so this study can ensure alignment of the working conditions of the new job with those of regular workers with stable roles and routines. Therefore, under the State Public Officials Act, which outlines the personnel regulations of public institutions in Korea, 22 regular workers who had completed the 6-month probationary period were selected as the standard representatives and enrolled as participants. During the interview process, theoretical sampling was conducted to thoroughly understand the differences in the subjects’ past work experience, level of education, and civil status. Having 22 participants is crucial to achieve theoretical sampling given how interview content was frequently based on the number of participants who shared similar characteristics as per previous studies [15, 22].

### Recruitment

Data collection was conducted through one-on-one in-depth interviews from April through August 2019. The study employed purposive sampling techniques to recruit participants by utilizing the authors’ personal network. The initial recruitment involved civil servants in the fire service, correctional civil servants, and an allied health worker. Subsequently, a snowball sampling method was utilized, wherein participants introduced other eligible participants, who were then recruited. In addition, members of the National Health Insurance Corporation—a prominent Korean public organization where many male nurses work—were also recruited after internal promotions and discussions of the study’s purpose and data collection procedures. Given how the number of male nurses working as school nurses was still relatively small at this time, the study also recruited members of the Korean Mannurses Association.

In-depth interviews were conducted until the theoretical saturation of the data contents was confirmed, wherein no new categories were found in analyzing the collected data. Following interviews with male nurses from various fields in the community, the study investigated the variations in adaptative experiences after transitioning to a community institution based on factors such as civil status, religion, and degree of education. To enhance the study’s comprehensiveness, additional participants were purposively recruited and interviewed.

Data collection through in-depth interviews was conducted by double-recording using cellular phones and tape recorders and included field notes and memos written by researchers in addition to the interview transcripts. The interviews were conducted at a café, in deference to the preferences of the research participants, and the interview times were determined according to their convenience. The semi-structured interviews utilized an open-ended format, starting with casual conversations aimed at establishing trust and creating a comfortable atmosphere between the researchers and participants before gradually progressing to the study’s topics. The initial questions were open-ended and broad to encourage participants to share as much as they can with regard to their experiences. As the interviews progressed, more specific questions were added as necessary (Table 1). Each interview lasted 60 to 90 minutes, and each participant was interviewed once or twice. In the data collection and analysis process, additional interviews were conducted if there was a concept that had not been confirmed by participants who had already been interviewed. After face-to-face interviews were conducted with the 22 participants, 4 face-to-face interviews and 4 additional telephone interviews were conducted for 8 participants for whom additional interviews were judged as necessary.

**Table 1.** Sample Interview Questions on Male Nurses’ Adaptative Experiences after Transition to Community Institutions.

| Category | Items |
| --- | --- |
| Main question | • Tell me about your adaptation experiences after moving to community institutions. |
| Supporting questions | • What did you feel in that situation?<br>• What were your thoughts in that situation? |

### Data analysis

Data analysis was conducted simultaneously with data collection, and open coding, axis coding, and selective coding were performed step by step according to the grounded theory method suggested by Corbin and Strauss [21]. After the interview, a researcher tried to complete the transcription work within 24 hours. However, given the island area where the study participants lived, transportation time was limited, so the interview content transcription of a few study participants took up to 48 hours. By repetitively reading the transcribed interviews, the field notes created during the interviews, and the memos, the data were analyzed line by line, concepts were classified, and similar concepts were organized into subcategories. In addition, the relationships between categories derived through open coding were organized into a paradigm model that includes causality, situation, phenomenon, intervening conditions, strategy, and outcome based on the dimension of categories and attributes. By thinking about the relationship between the concepts and subcategories obtained through the analysis, the core category was derived, and propositions were derived through relational statements, thus leading to a substantial theory that could explain the male nurses’ adaptation experiences after moving to community institutions.

### Researcher preparation

Before this study was initiated, the researchers had experience working with male nurses in general wards, special units, and intensive care units of tertiary general hospitals. Additionally, as university lecturers and through student career counseling, the investigators have learned that male students in nursing colleges experience worry about their careers. Moreover, the investigators found that male nurses employed in clinical roles were worried about adapting to the clinical sector and changing jobs. We, therefore, became interested in male nurses changing jobs to those in community institutions. Our practical and educational experiences helped us to understand the research participants’ situations holistically. Not only have we conducted numerous qualitative studies and published many papers, but we have also continuously participated in qualitative research–related conferences and seminars to exchange opinions with other scholars studying grounded theory, and we have made continuous efforts to improve the quality and sensitivity of related research.

### Study validity and rigor

This study was validated by applying the four-tiered criteria of reliability, applicability, consistency, and neutrality, as suggested by Guba and Lincoln [23]. First, reliability is the concept of whether the research results accurately measure reality. We directly conducted semi-structured interviews using open-ended questions, and we repeatedly checked and transcribed the directly recorded contents so that no interview data were omitted. Additionally, efforts were made to ensure reliability by continuously comparing the transcribed data and the results derived from the analysis process and repeatedly checking whether the contents matched. Furthermore, we collected data with sensitivity to the research participants to increase the reliability of the research. The results were confirmed, corrected, and supplemented by five nursing professors with rich experience in qualitative research. Second, applicability refers to whether the research results can be applied in similar situations other than the one in which the research was conducted and whether readers can accept the research results meaningfully in light of their own experiences. The study was presented to two male nurses with experience in job transfers to local community organizations and who had not participated in the study. Additionally, two researchers with qualitative research experience who did not participate in this study presented the research results and received advice to maximize the applicability of the research results. Third, to ensure consistency or whether the same results can be obtained if the study is repeated, we described the analysis process in detail during data collection and strictly following the study procedures. Five nursing professors with rich experience in qualitative research were asked to evaluate the analysis process. Fourth, neutrality ensures the objectivity of the research process and results. A literature review was conducted after data collection and analysis were completed to a certain extent such that the contents of the literature review did not prejudice our conduct of the study procedures and analysis. Additionally, the participants were asked multiple times to confirm their understanding so that their experiences were captured as fully as possible and the researchers’ past experiences did not interfere with the study.

### Ethical consideration

This study was approved by the Chonnam National University Bioethics Review Committee (IRB No. 1040198-180720-HR-070-03). The purpose and methods of the study were explained to each study participant, and all participants expressed their intention to participate and provided written consent. The participants were assured they could pause or end interviews at any time, and the interview durations were communicated beforehand. Moreover, the researchers explained that the entire interview would be recorded, that the recorded and transcribed data would be used only for research purposes, that the research data would be safely discarded immediately after the storage period after the completion of the research, and that the anonymity of the research participants was guaranteed.

## Results

### General characteristics of participants

The participants were in the age group of 29 to 39 years (mean 32.59 years); 11 participants were married, and 11 were unmarried. The participants had academic backgrounds ranging from junior college graduates to graduate school, and all participants were military veterans. They worked in various occupations, including as civil servants. All had worked at medical institutions for at least six months but not more than six years. At the time of enrollment into the study, eight participants worked at public institutions, eight were firefighters, four were correctional workers, one was a health-care worker, and one was an allied health worker. The ages at which they left their previous jobs ranged from 26 to 33 years (mean, 28.41 years). All participants had worked at their current jobs for at least seven months but not more than nine years (Table 2).

**Table 2.**
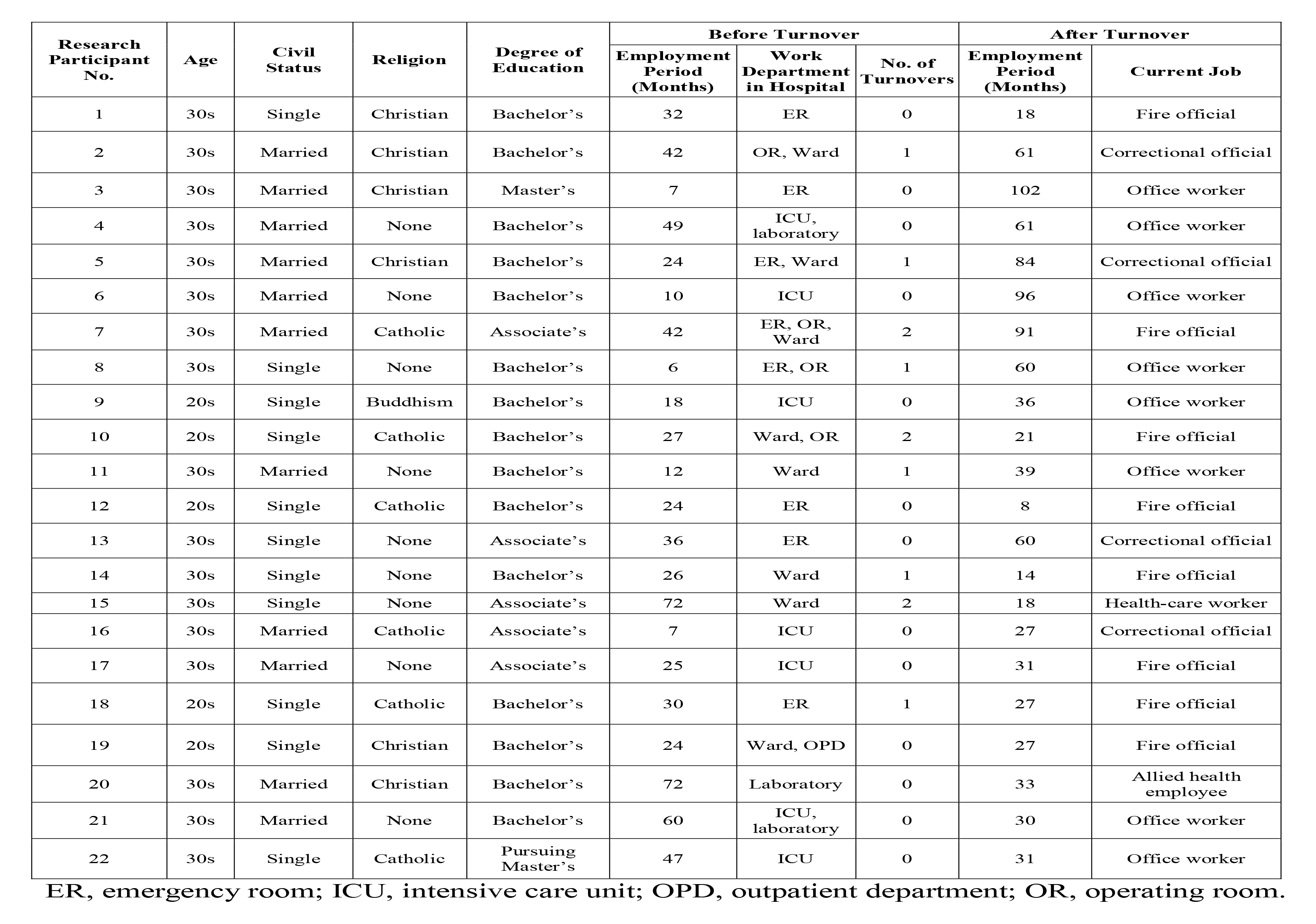
General Characteristics of Participants.

### Adjustment experiences of male nurses after moving to community institutions

The analysis of the data on the adjustment experiences of male nurses after moving to community institutions, with categorization of the interview data through open coding, yielded 29 subcategories from the 11 main categories (Table 3). The categories were established through axial coding according to the paradigm model, and a situation model that integrated and explained all categories was presented (Fig 1). The core category of this study was “*putting down roots in another place for myself*.”

**Fig 1.**
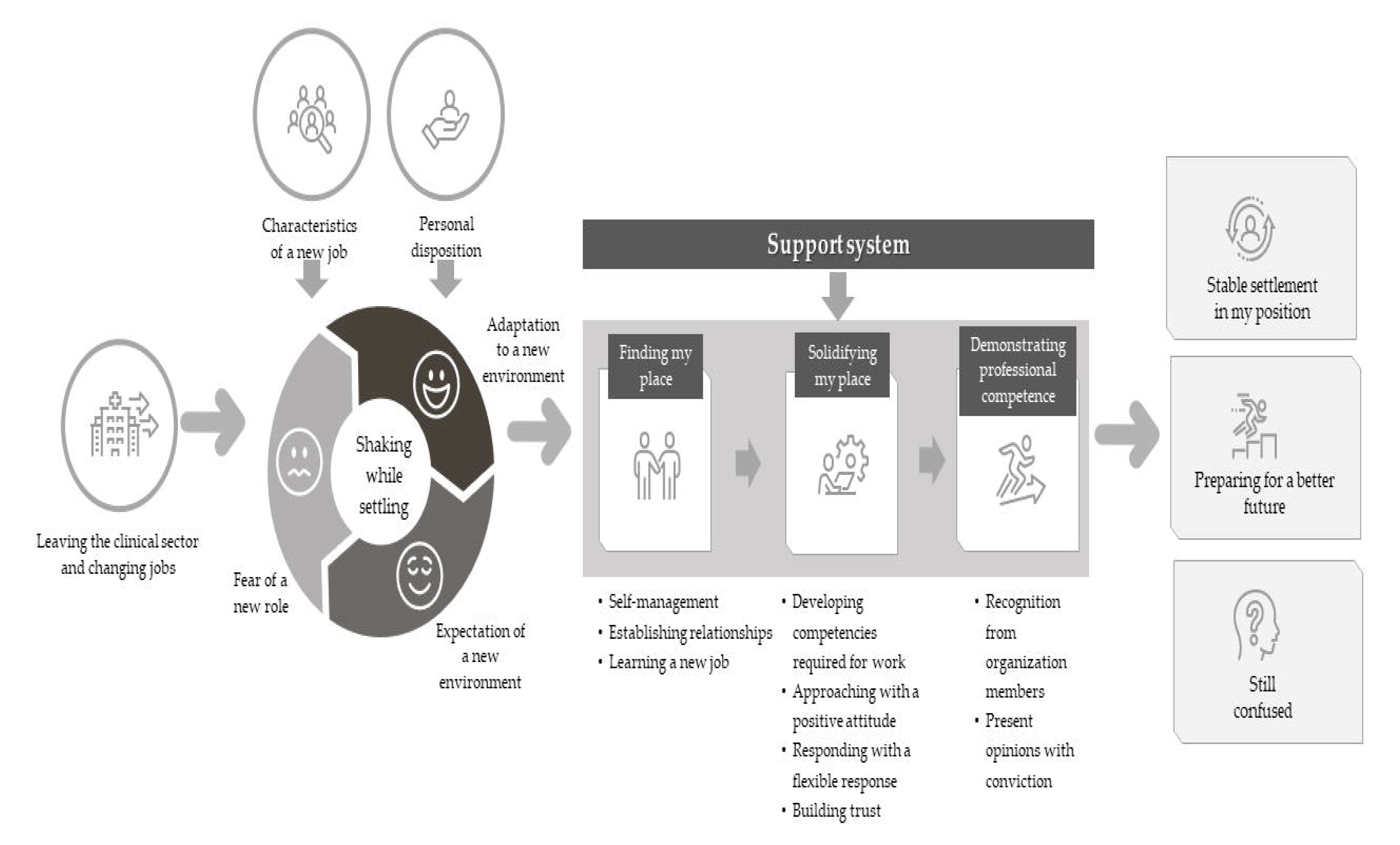
Male Nurses’ Adaptation Experiences after Turnover to Community Institutions in Korea.

**Table 3.**
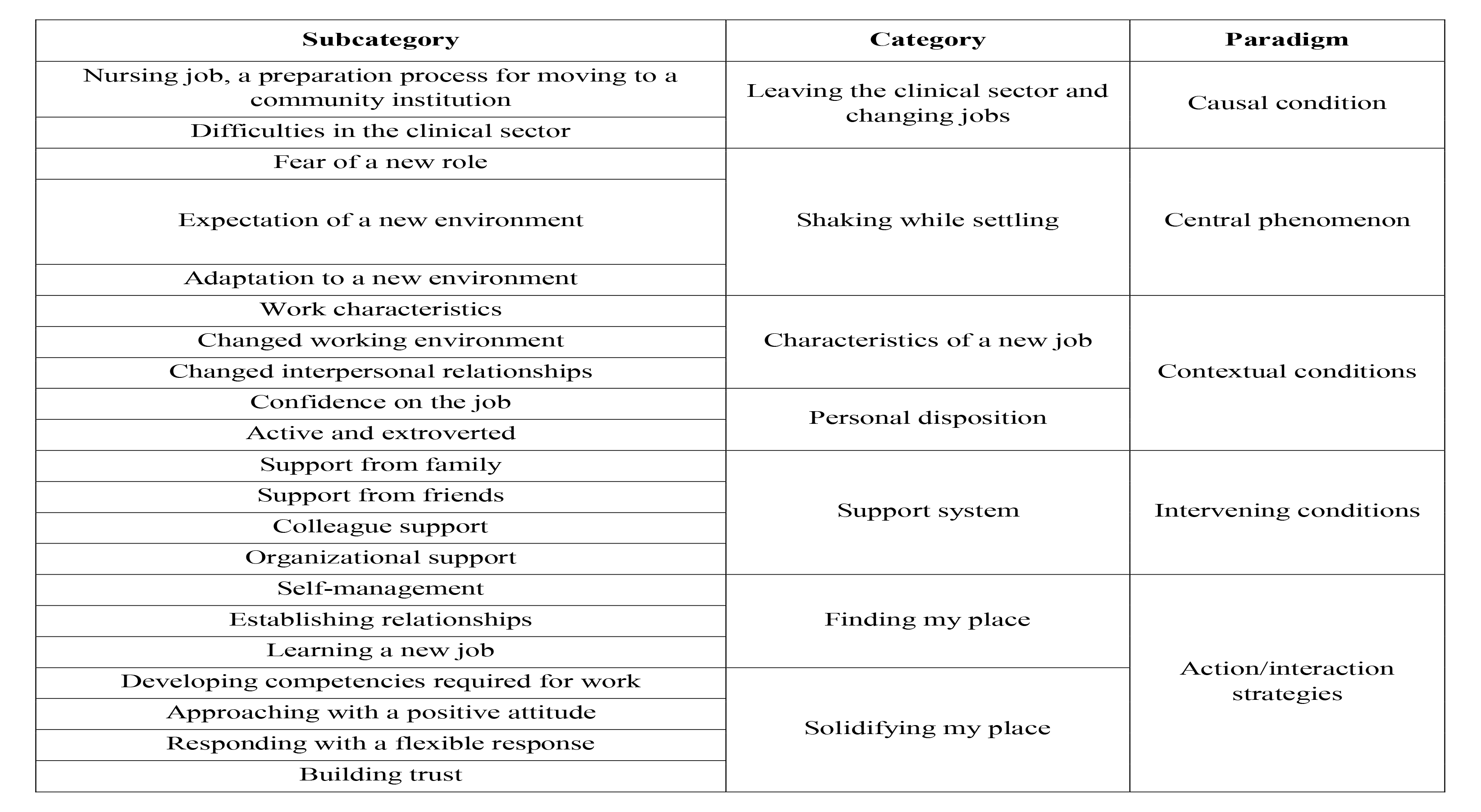
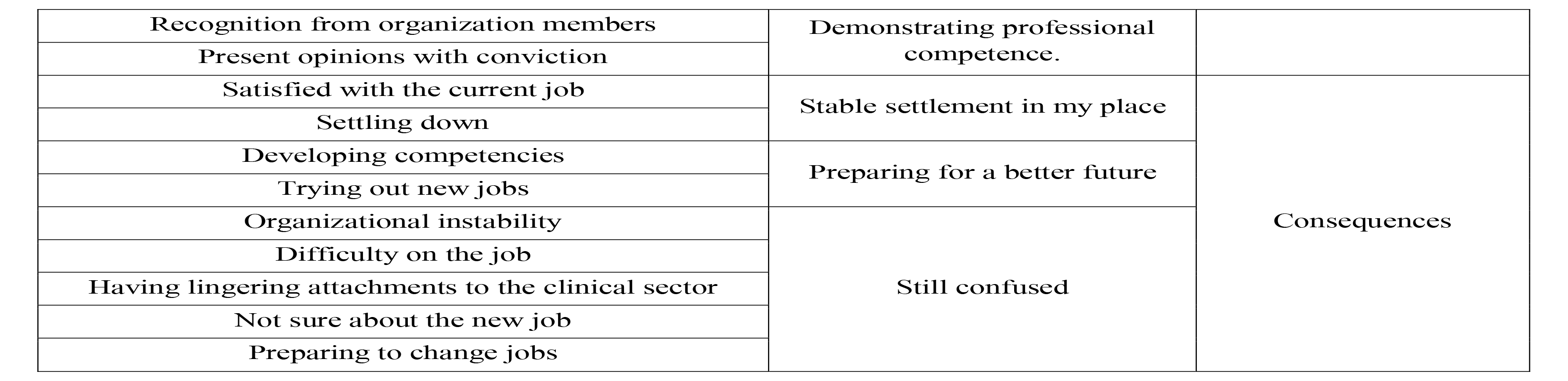
Paradigm, Categories, and Subcategories of Male Nurses’ Adaptation Experiences after Turnover to Community Institutions in Korea.

### Causal condition

A causal condition refers to a cause of a certain phenomenon [21], and in this study, the causal condition was *leaving the clinical sector and changing jobs*. The subcategories were *nursing job, a preparation process for moving to a community institution,* and *difficulties in the clinical sector.* Some research participants considered clinical nursing a means of transitioning to other occupations from the beginning, so they achieved the basic clinical sector requirements and moved to community institutions. A minority of the study participants left the clinical sector and moved to community organizations with the personal belief that they wanted to work in a field they were more interested in rather than continuing negative experiences in the clinical sector. Male nurses who chose the nursing profession because of the ease of employment experienced various difficulties in the clinical sector after joining hospitals as new nurses. In the process of adapting to clinical work, they experienced dissatisfaction with their working environments, difficulties forming human relationships, social prejudices against male nurses, and difficulties, with their gender role of being responsible for the livelihoods of their families. In the end, the male nurses, who had expressed their worries, moved to local community organizations through the process of preparing for a job change.

- “I found out about fire service paramedics when I was a college student. To apply as a firefighting paramedic, I needed two years of clinical experience. So, I thought I would work at the hospital unconditionally for two years” (Participant 12).
- “At the hospital, the nurses were always busy and had a lot of work, so they were frowning. Today and tomorrow, I thought that the loading of this work would not change. In the hospital, I could often see nurses being scolded by senior nurses. In particular, new nurses were scolded by senior nurses even in front of patients, and they seemed to have no human rights. So, I thought of going to the medical insurance review and assessment service or the health insurance corporation after completing my clinical career at a hospital” (Participant 2).
- “I think the public awareness about nurses is still a bit low. Older adults still think negatively about male nurses working in hospitals” (Participant 4).
- “I’ve heard this from patients: ‘Why is a man here? Call her.’ When I was treated like this by a patient, my self-esteem was very low” (Participant 18).
- “I’ve never seen senior male nurses working until the age of 40 or 50. So, I always thought about it” (Participant 19).

### Central phenomenon

A central phenomenon is an answer to the question, “What is going on here?” This is under the influence of causal conditions, referring to a central idea or event occurring in the phenomenon [21]. The response indicates how the participants in the study take action on the problem or situation they are facing, and it is controlled by strategic action or interaction. In this study, the central phenomenon experienced by the research participants was shaking while settling. The subcategories were *fear of a new role, expectation of a new environment,* and *adaptation to a new environment*.

As the study participants left the clinical sector and moved to community institutions, they initially worried about the new job, were concerned about the eyes of the people around them, and experienced fear of their new roles while experiencing nervousness about mistakes. Furthermore, they experienced the expectations of themselves and the people around them in the new environment. This experience caused various conflict situations and made the participants experience major and minor anxiety. However, the research participants focused on new work and accomplished tasks while feeling pleasure, focused on forming relationships with members of the organizations, experienced comfortable relationships in their new organizations, and tried to settle down while gradually adapting to their new workplaces.

### Contextual condition

Context refers to a set of specific circumstances, factors, or situations that contribute to a central phenomenon or problem [21]. In this study, *characteristics of a new job* and *personal disposition* appeared as contextual conditions. The subcategories of *characteristics of a new job* were *work characteristics, changed working environment,* and *changed interpersonal relationships*. The subcategories of *personal disposition* were *confidence on the job* and *active and extroverted*.

The participants felt burdened and stressed because of the tasks that differed from what they were used to performing as clinical nurses. Although they were hired as special hires based on their nursing qualifications and experience, in the case of firefighting civil servants, the work burden was high as they additionally performed tasks, such as security, rescue, and fire suppression, that had nothing to do with their backgrounds. The participants, who had been physically tired of irregular shift work and night shifts in the clinical sector, were satisfied with the absence of night shifts at their new workplaces. Although some of the participants were working longer hours because of double shifts, it was found that they did not suffer greatly from the extended working hours because of the allotment of breaks. Some participants still had to work shifts similar to those in hospitals, but they did not feel much discomfort with the change in work patterns because the work patterns were not as complex or irregular as in hospitals. Additionally, some participants who worked at public institutions said that business trips were repeated frequently and felt uncomfortable. By performing new tasks related to their educational and previous professional backgrounds, the research participants could feel professional pride because their experiences as nurses had greatly helped them resolve their current tasks, and their expertise was recognized. Correctional civil servants felt uncomfortable because they were isolated from the outside world during working hours because of the environmental specificity of prison, and they experienced a negative social gaze.

- “Ordinary prison guards only have to work as prison guards, but as I am both a prison guard and a nurse, I work as both a prison guard and a nurse” (Participant 16).
- “My job is to go to the scene by ambulance, address the situation or treat the patients, and transport the patient to the hospital. It’s just that it’s not a clinical setting, but here, too, we do work related to nursing. There are emergency medical technicians, but nurses are more accurate in looking at patients and do a better job” (Participant 7).
- “Because it is a group of men rather than women, there were no difficulties from gender or discomfort in human relationships like in the hospital. Women are a bit talkative, but it was a little more comfortable here” (Participant 10).
- “I’m satisfied when I’m in prison, but one regrettable thing is because of social awareness. Public awareness remains low. It will probably be like that until I retire” (Participant 5).

In terms of personal characteristics, some of the research participants had high self-esteem, expressing their convictions about their jobs. On the one hand, participants with receptive, positive, and active personalities were more easily adaptable to their new organizations. On the other hand, participants with meticulous personalities felt stress and difficulty at the beginning because of this personality during work, but after adapting to the work, this personality became an advantage.

- “I can help others with something, and there is a sense of reward from work.” (Participant 3).
- “I’m a little meticulous. I think it was a bit stressful because there was something telling me that I shouldn’t leave out something.” (Participant 10).
- “My original personality is a little bit individualistic. I hate getting entangled with others. I don’t like helping or receiving help from others. I don’t like engaging and being harmed in the first place. Silently doing what I have to do.” (Participant 18).

### Intervening condition

An intervening condition is a condition that alleviates the intensity of a phenomenon experienced by a participant or induces a change—a factor that promotes or hinders actions or interactions [21]. The *support system* was found to be an intervening condition that affects the intensification or alleviation of the shaking while settling phenomenon. The subcategories of *support system* were *support from family, support from friends, colleague support,* and *organizational support*. When the participants experienced anxiety (shaking) in the process of adjusting after changing jobs, their wives, parents, and friends helped them adapt, and their seniors, team members, and superiors helped them adapt via advice, praise, and encouragement. Additionally, research participants who worked in organizations with standardized work guidelines and systematic training support programs adapted to their new organizations more easily.

- “When I was having a hard time, my wife helped me a lot. She told me a lot that I could do well” (Participant 17).
- “First of all, motivation was the most important. I asked a lot of my classmates. As we are all in the same position, it may have been a little rude to ask. Still, as I was motivated to do the same job, I asked a lot and got help” (Participant 4).
- “I think the biggest advantage is that there are manuals and instructions. At the hospital, each person has a different reaction, progress, and individual differences, but office work doesn’t lie. It’s clear what results come out when you check something. I liked it because there seemed to be less uncertainty” (Participant 9).
- “We have a 1-year mentoring program, so four to six mentees go into one mentor group and tell us a lot about the company life know-how and general stories. Additionally, the work is thoroughly divided here, so I go to the person who corresponds to the work and learn the work” (Participant 20).

### Action/interaction strategies

In the grounded theory method, action/interaction refers to strategies, routines, and habits to cope with problems in a conceptual way that answers the questions of “who” and “how” [21]. The actions/interactions shown in this study were *finding my place, solidifying my place,* and *demonstrating professional competence*. The subcategories of *finding my place* were *self-management, establishing relationships,* and *learning a new job*. The subcategories of *solidifying my place* were *developing competencies required for work, approaching with a positive attitude, responding with a flexible response*, and *building trust.* The subcategories of *demonstrating professional competence* were *recognition from organization members* and *present opinions with conviction*.

The participants had interactions to adapt to new jobs and find their places. During this time, research participants tried to control their emotions through self-management. The participants compared their past clinical sector experiences, found the strengths of their current jobs, and tried to be more satisfied. Additionally, to better perform the roles required within the new organizations and interpersonal relationships, the participants valued the relationships between members of their organizations and specified behaviors for organizational adaptation. They assumed a receptive stance, adjusting their habits to fit the habits, roles, requirements, and preferences of those around them, accepting suggestions from colleagues and following organizational guidelines. Furthermore, mind-opening strategies were used, such as expressing gratitude, acting politely, and acting proactively.

- “I think I sorted it out while thinking about my daily routine before I went home after work and went to bed. If you feel that you spoke too quickly when you were talking to this person today, go the next day and spend a little more time with that person and adjust your speaking speed” (Participant 20).
- “To develop a rapport, we drank coffee together, talked about personal things, and tried to get closer to each other” (Participant 17).

After the participants found their places in their new environments, they made efforts to secure these places. They worked hard to further develop the competencies necessary for their jobs and worked with positive attitudes. They tried to build deep trust beyond the formation of relationships with others, and they made efforts to deal resolutely even in uncomfortable situations.

- “When you live with other people, things that are a little sad or regretful happen. Then, I think I took control of myself by making up my mind to think positively” (Participant 1).
- “Living as a person is not always about doing more work. Depending on the situation, I think I need some flexibility in my life” (Participant 16).

Study participants gradually settled into stable positions in their new jobs over time. Additionally, they did not settle for the present but exerted efforts for self-development while making academic efforts, and they demonstrated their professional capabilities by actively presenting their opinions in their specialized fields and revealing their convictions.

- “I have to write a lot of reports here, but I felt that I was lacking in grammar or systematic writing skills. So, I bought several books separately and studied them, and I am still studying them” (Participant 20).
- “In situations where all employees feel unreasonable, I have my say” (Participant 13).

### Consequences

In this study, the three categories of “stable settlement in my place,” “preparing for a better future,” and “still confused” were derived through the process of action/interaction of the participants. The subcategories of *stable settlement in my place* were *satisfied with the current job and settling down*, and the subcategories of *preparing for a better future* were *developing competencies* and *trying out new jobs*. The subcategories of *still confused* were *organizational instability, difficulty on the job, having lingering attachments to the clinical sector, not sure about the new job,* and *preparing to change jobs*.

As a result of the adaptation process after changing jobs, the participants showed satisfaction and settlement in their current workplaces. The adaptation process after changing jobs was not easy; however, the tension and psychological burden were lower when compared with the clinical sector, and it was less physically demanding because there was not as much work. In addition, they liked the salary and welfare system. They were able to have time to spare as they established a work-life balance, and they liked the promotion system. For these various reasons, the research participants expressed satisfaction with their current jobs and settled down in reality as they thought that their current jobs were their lifelong jobs moving forward.

- “Male employees are also taking parental leave these days. There are actually employees who use it, and you can use it freely without notice” (Participant 8).
- “Nurses have to follow a set pattern, and there are many difficult parts to adjust to working there, but the public servants let me rest at the time I want. I think it’s much more comfortable because I have more free time” (Participant 5).

Participants showed how they prepared for a better future by developing their capabilities and changing jobs in the adaptation process after changing jobs. To develop their competency, they made plans to go to graduate school, get master’s degrees, and study their fields of interest. To develop the skills required for work, they continued to study and be prepared for opportunities for promotion. They changed their occupations and took on new jobs more aligned with their inclinations.

- “I have a lot of foreign business trips, but speaking and listening in English is not enough. So, I go to private academies and continue to study English over the phone” (Participant 20).

Not all participants were satisfied with the results of the adaptation process after changing jobs. Some participants still seemed confused about their jobs. The causes of the confusion included the instability of the organizations they moved to, job stress, and lack of confidence in the job. Additionally, attachments to the clinical sector remained. However, the participants did not dream of moving to other jobs right away, and after working for another 1–2 years, they said they would consider moving to new jobs. However, some participants felt they could no longer get a sense of accomplishment from the new job and were actively preparing for job changes.

- “I wonder if this is definitely my job. I’m not sure yet. I think I might change jobs if I find a more stable job” (Participant 18).

## Discussion

This qualitative study applied the grounded theory method to explore male nurses’ adjustment experiences after moving to community institutions and to develop a substantive theory accordingly. In the results of this study, the central phenomenon experienced by male nurses in the adaptation process after moving to a community institution was shaking while settling, and the core category was “putting down roots in another place for myself.” The following is a detailed look at the relationship between the conditions that appeared in the male nurses’ adaptation experiences after moving to community institutions.

The core category of this study refers to a series of processes in which male nurses cope with and adapt to the phenomena they experience while moving to community institutions. In previous studies, male nurses’ turnover experiences showed that male nurses moved from one clinical field to another to find stable positions for themselves [24]. In contrast, in this study, male nurses who experienced conflict and confusion in the clinical sector moved to community institutions that recognized their positions as more stable, and they settled down while gaining professional expertise and forming a professional identity in other occupational settings. The study showed how they developed, took root, and settled down while becoming accustomed to their new roles.

On evaluating the causal condition of leaving the clinical sector and changing jobs, we found that male nurses who chose nursing because of the ease of employment advanced to other fields rather than working as nurses for a long time, which is consistent with the findings of previous studies [25]. Nursing was recognized as an intermediate bridge until the completion of two years of clinical service, which is a requirement for employment as a civil servant firefighter. Therefore, it has been considered that a customized curriculum is needed to cultivate male nurses’ nursing professionalism and to explore various careers within the nursing sphere. It was found that the research participants changed their jobs in search of good working conditions to receive proper treatment and compensation as opposed to unsatisfactory wages, welfare conditions, and unwanted department work. This result is similar to that of previous studies, which reported that working conditions, such as satisfaction with the current workplace and salary level, affect nurses’ intention to leave [23,25,26]. Currently, South Korea does not have a wage guideline for nurses and relies on the wage system for each institution; however, efforts to establish a standardized wage guideline for nurses have been proposed. Some participants had difficulties in relationships with female nurses and felt they were not treated with due respect, resulting in low self-esteem and difficulties in vertical relationships with doctors. Additionally, some participants were disappointed with their managers’ attitudes and had difficulties in personal relationships because they lacked supportive fellow nurses. This was similar to a previous study that found that male nurses felt alienated from female-centered nursing organizations and had difficulty forming relationships and communicating with female nurses, resulting in reduced job satisfaction and high turnover [6]. Therefore, it is necessary to develop an emotional communication education program in which male nurses are relatively lacking to make efforts to understand each other’s differences and maintain good interpersonal relationships with colleagues. Some of the study participants left the clinical sector because of difficulties that they experienced as men, such as gender role stereotypes, the burden of raising a family, the absence of male nurse role models, and difficulties with performing multiple tasks at the same time. Social prejudice against male nurses exists, and patients who are unfamiliar with being nursed by males may have negative reactions [1,28]. The public perception that nursing is a traditionally female-dominated profession is an obstacle to male nurses’ recruitment and professional development [29]. Male nurses are still excluded from promotion within some organizations, and the absence of role models remains a problem [5,30]. It is necessary to establish a management system in which opportunities for promotion and compensation are fair, considering the clinical careers of male nurses, and to discover cases of male nurses promoted to managerial positions (such as head nurses), share their stories, and present them as role models. To optimize the efficient work performance of male nurses, we propose a method of providing educational management guidelines while diversifying the preceptor training period and content according to the clinical adaptation skill level of the male nurse. The guideline should precede the development of hospital practical education programs for male nurses, such as assigning nurses exclusively in charge of education of the same sex, securing the required training period, and preparing a customized education curriculum, considering the hospital context.

The contextual conditions that generally affect the adaptative process of male nurses after moving to community institutions in this study included the characteristics of a new job and personal disposition. Research participants experienced job stress because of the work burden in the early days of joining new companies. Away from the clinical sector, new working contexts, such as firefighting, correctional work, public institutions, schools, and pharmaceutical companies, were new areas of expansion for male nurses, and at the same time, a life event associated with apprehension (shaking) while trying to settle. Therefore, a strategy to develop and utilize various educational tools is proposed, considering the job characteristics of the participants so that male nurses working in various occupations can use the currently implemented 8-hour supplementary education for nurses annually. Additionally, it will be necessary to develop a step-by-step supplementary education system suitable for male nurses in each job field and establish an application strategy to be an educational system for professional development.

Among the characteristics of new jobs, the shift from three-shift work to full-time and two-shift work increased job satisfaction, which was consistent with the results of a previous study [31], which associated non-shift work with a higher quality of life than shift work. Nurses experience physical exhaustion and reduced quality of life because of night shifts and irregular shift work. Therefore, from the stage of preparing the work schedule, shift work should be adjusted as regularly as possible with considerations of biorhythms, expanding the level of support for night nurses to improve working environments for male nurses, guaranteeing work options and health rights, as well as desirable work patterns. A national-level policy for model development is required.

In contrast, civil servants in correctional positions experienced job stress because of the special working environment of prisons and negative social perceptions, which was consistent with the results of previous studies [15,32,33]. Nursing in correctional facilities is thought to require the operation of programs for stress management for correctional nurses, considering that hospitals, community health centers, and industries have different organizations and working environments, and the subjects to be cared for and the terms of service vary. Additionally, health teachers felt a difference in the social perception of health teachers compared with other teachers. This was similar to the results of a previous study [20], as elementary school health teachers felt a sense of occupational presence and confusion about occupational identity. While having two qualifications, nurse and teacher, health teachers experience confusion in their professional identity. They are affected by interpersonal relationships with colleagues, public perceptions of their jobs, and their work or role, and they adapt to the role of health teachers. Policy support against social inequality can help individuals provide nursing care and live a life as a teacher; this can include support for class research for professional development and expansion of opportunities for class-related job training. Additionally, policy support and cooperation among nursing organizations are necessary to continue the improvement of the health teacher promotion systems and performance-based pay systems to increase the number of health teachers in large classes and to improve the status of health teachers.

The participants experienced changes in interpersonal relationships as the number of male colleagues increased, and they formed positive peer relationships in organizations because of the changed nature of their jobs. As the number of same-sex colleagues increased, candid conversations became more possible, and difficulties and discomfort in human relationships because of gender were eliminated. In a previous study, when satisfaction with human relations was high, work tenures were prolonged, and positive human relationships played an important role in the path between dissatisfaction with a company and intention to leave the job [35]. Male nurses are alienated from female-centered nursing organizations and have difficulty forming relationships and communicating with female nurses, resulting in reduced job satisfaction and high turnover [5]. Therefore, it is necessary to make individual efforts to understand the differences between men and women. One proposal in this regard is to establish a gender equality curriculum as a liberal arts subject in nursing college curricula.

With regard to personal disposition, people with receptive personalities, positive personalities, and active personalities found it easy to adapt to transitions to community organizations, and meticulous personalities found it difficult in the early stages. However, associated stress dissipated after adjusting to new job performance requirements and routines. People with individualistic personalities, who silently do what they have to do, may find themselves in a state of confusion when adapting to work life. In a previous study [35], nurses with personality type D, who showed a tendency to be vulnerable to negative emotions and suppressed self-expression in social interactions, showed significantly lower job satisfaction and nursing professionalism, which was similar to this study. Another study investigating male nurses [36] found that people with more sociable personalities used more active coping methods, such as collecting information, making plans, and solving problems, than people of the “stable type,” which was similar to the results of this study. Therefore, to address anxiety (shaking) among workers adapting to a new environment after changing jobs, workplaces can consider developing stress-coping education programs with consideration of personality types.

The examination of the interventional conditions that strengthen or weaken the action/interaction shown in this study is a crucial process for developing interventional strategies to help male nurses adapt in transitions to community institutions from a nursing perspective. In this study, the mediation condition was the support system, which included support from family and friends, peer support, and organizational support. Wives, parents, and friends acted as supportive resources, whereas advice from colleagues, help from seniors, support and help from team members, and praise and encouragement from superiors acted as supportive resources. Standardized work guidelines or systematic training programs could also be used as organizational support. Among various social supports, family support, in particular, strengthens an individual’s capacity to adapt to problems in stressful situations [37], and the family, as the primary support system, plays a role in forming self-esteem and increasing life satisfaction as the degree of support increases. [38]. Therefore, family support is considered an essential factor in facilitating coping with crises and adapting to change [39]. Additionally, in the results of this study, support from supervisors and peers affected male nurses’ job satisfaction [30,40], and in previous studies, job satisfaction was influenced by a supportive atmosphere and systematic education programs to attract new nurses and help them adapt. In general, support has been shown to be an important factor in recruiting and retaining employees [41]. It is necessary to systematically provide opportunities by preparing human relationship improvement programs so that support systems, which are positive mediation conditions that strengthen interaction, can be sufficiently utilized [42]. Based on horizontal communication with managers, it is necessary to prepare and continuously manage managerial leadership promotion education so that the ability of conversational leadership or coaching leadership can be cultivated.

Even in light of the central phenomenon of shaking while settling, male nurses working in community organizations used various interaction strategies. According to the theory of work adjustment of Dawis and Lofquist [43], when an individual becomes dissatisfied, he or she actively tries to change the environment or adapts through passive efforts to change himself or herself. This study found that the participants used passive strategies to change themselves at the beginning, and after a certain period, they used active strategies, such as self-development and revealing their convictions. In a previous study [44,45], paying attention to one’s surroundings, taking time to participate in private meetings, and expressing sympathy for other people’s words were indicators of adaptation while actively participating in meetings to maintain good relationships with female nurses. This was concordant with the results of the present study. Additionally, the pattern of adapting one’s habits as one adjusts is consistent with previous studies; this refers to identifying other people’s tendencies and acting accordingly rather than expecting other people to change [44,45].

Some participants showed stable settlement in their positions or prepared for a better future, but a minority were still confused. In particular, correctional nurses experienced much role conflict. A previous study investigated correctional nurses performing roles, such as drug preparation and treatment, when there was a shortage of medical personnel because of the absence of pharmacists and medical officers [19]. Licensed and registered male nurses were recruited, but they experienced role confusion as they worked in the general security field instead of working only in the medical field. Therefore, clear institutional standards for post-employment work should be established to enhance male correctional nurses’ professionalism and role identity. Additionally, the eligibility requirements for the special recruitment of correctional workers were stipulated to be characteristics or skills of mental health nurses or nurse license holders, and about 62% of the correctional facility’s occupancy was classified as patients, and mental illness, hypertension, and diabetes accounted for about 74% of all diseases [46]. To manage the health problems of prisoners to a certain level, better nursing care would be provided if nurses with various experiences in clinical practice were employed. Unlike clinical or community nursing, nurses in correctional facilities have the additional responsibility of facilitating correction or forensic rehabilitation. Therefore, specialized and customized education should be provided through job development that reflects the characteristics of the medical field and correctional facility, and an environment in which nursing practices should be established—with reciprocal support structures and recognized expertise—should be created by activating internal exchanges.

Clark divided the roles of community nurses into patient-centered roles, delivery-centered roles, and population-centered roles, and suggested the roles of direct nursing providers, educators, counselors, referral resources, role models, and case managers as the patient-centered roles [47]. However, another preceding study reported that community nursing is ambiguous, difficult to associate, and confusing compared with direct nursing interventions in clinical settings, particularly because community nursing is conceptual and abstract as it is learned as theory in the classroom. In the United States, a group called the Henry Street Consortium was formed to facilitate the work of community nurses, and through related literature, actual cases, and interviews, the core competencies required for health nurses to perform their duties in the field were comprehensively reflected. A Public Health Clinical Manual was completed [48]. This manual organizes 11 core competencies of community nurses and cites actual cases where these competencies are implemented so that even students who have no knowledge of health nursing or new health nurses can understand the details of their job descriptions [48]. To nurture nurses needed by the community, it is necessary to ensure that practical training in community nursing is designed in various ways based on the core competencies required of actual community nurses and operated to facilitate competency acquisition naturally.

There are limitations in generalizing the results to all male nurses. As only volunteers participated in this study, it is possible that male nurses who were reluctant to share their stories or those who had closed personalities were omitted. Therefore, care should be taken not to generalize our findings to all male nurses. This research focused on community organizations where many male nurses work, and we tried to select research participants from various community organizations using snowball sampling; however, there exists a possibility that male nurses from particular community organizations were omitted.

## Conclusion

In this qualitative study wherein the grounded theory method was applied to explore male nurses’ adaptative experiences after moving to community institutions and to develop a substantive theory that can explain the relationship between various complex and related conditions, we found that the adaptative experiences vary by the degree of activeness in undertaking interactive strategies and utilizing multiple support resources. This suggests that active organizational support is necessary so that people around transitioned nurses can actively intervene and utilize various support resources in the flow of time, going through trial and error in the acceptor phase and going through the concentration phase, and the stabilization and leap phase, with their own efforts. The results of this study can help male nurses understand the adaptation experiences associated with leaving clinical institutions for community institutions, identify problems experienced in the process of adaptation after leaving a job, and develop programs to seek strategies to support stable adaptation to a new job. It is meaningful in providing useful basic data for development. This study suggests the need for active help and policy support from nursing institutions to ensure that male nurses can expand the scope of nursing into a new environment of community institutions while maintaining the identity and professionalism of nurses. Based on the reality theory of this study, it is necessary to develop and verify the effectiveness of a program that can help male nurses adapt by considering various conditions that affect their adaptation after switching to community institutions.

## Author contributions

Conceptualization, JK and HCC; data collection: JK; formal analysis, JK, SK, and HCC; writing – original draft preparation, JK and SK; supervision, HCC; project administration, JK; writing – review and editing, SK and HCC. All authors have read and agreed to the published version of the manuscript.

## Supporting information

IRB Certificate of Approval

Clinical Studies Checklist

## Data Availability

Data cannot be shared publicly because of privacy requirements.

## Acknowledgments

The authors would like to express their heartfelt appreciation to the participants of this study, as this study would not have been possible without their valuable time and support.

## References

1. Kim JH, Park KO, Kim JK; Yun HJ, Lee JH, Cho EK, Kim SH, Kim YH. An adaptation experience of male nurses at general nursing unit. J. Korean Acad. Nurs. Adm. 2016;22(5):496–506. doi: 10.11111/jkana.2016.22.5.496

2. Lee JS. The era of 20,000 male nurses has opened. The Korean Nurse Association News [cited by 2022 Sep 20]. Available from: http://www.nursenews.co.kr/main/ArticleDetailView.asp?sSection=61&idx=24940&intPage=5

3. Kim IJ, Shim HW. Subjectivity about turnover intention among male nurses in South Korea: A Q-methodological study. Asian Nurs Res. 2018;12(2):113–120. doi: 10.1016/j.anr.2018.04.002

4. Cho MK, Kim CG, Mo HJ. Influence of interpersonal relation and job stress on nursing performance of male Nurses. J Muscle Joint Health. 2015;22(3):195–204. doi: 10.5953/JMJH.2015.22.3.195

5. Zhang H, Tu J. The working experiences of male nurses in China: Implications for male nurse recruitment and retention. J Nurs Manag. 2020;28(2):441–449. doi: 10.1111/jonm.12950

6. Ashkenazi L, Livshiz-Riven I, Romem P, Grinstein-Cohen O. Male nurses in Israel: Barriers, motivation, and how they are perceived by nursing students. J Prof Nurs 2017;33(2):162–169. doi: 10.1016/j.profnurs.2016.08.001

7. Choi KH, Kim HJ, Kim JH, Nam ES, Han HJ, Kang H, et al. Male nurses’ experiences of being rejected in nursing practice. J Korean Acad Soc Nurs Educ. 2018;24(1):16–28. doi: 10.5977/jkasne.2018.24.1.16

8. Oh S, Jang HY. A qualitative meta-synthesis on male nurses’ adaptation experiences as clinical nurses. J Korean Assoc Qual Res 2018;3:12–19. doi: 10.48000/KAQRKR.2018.12

9. Korean Nurses Association. Status of active nurses compared to licensed registered nurses [cited 2021 Jul 28]. Available from: http://www.koreanurse.or.kr/resources/statistics.php

10. Hospital Nurses Association. Investigation of the current status of hospital nursing staff placement [cited 2021 Jul 28]. Available from: https://khna.or.kr/home/pds/utilities.php

11. Shin JH, Seo MH, Lee MI. Nursing jobs and gender in our age of convergence: Research on male nurses. J Digit Converg. 2016;14(3):287–297. doi: 10.14400/JDC.2016.14.3.287

12. Diño MJS, Ong IL. Research, technology, education & scholarship in the fourth industrial revolution [4IR]: Influences in nursing and the health sciences. J Med Invest. 2019;66(1.2):3–7. doi: 10.2152/jmi.66.3

13. Wakefield M, Williams DR, Le Menestrel S. The future of nursing 2020-2030: Charting a path to achieve health equity. Washington, DC, USA: National Academy of Sciences; 2021.

14. Chin YR, Kim H. The role of community health nurse in assay written by a nurse practitioner of primary health care post. J Korean Public Health Nurs. 2016;30(2):300–310. doi: 10.5932/JKPHN.2016.30.2.300

15. Mao A, Cheong PL, Van IK, Tam, HL “I am called girl, but that doesn’t matter” - perspectives of male nurses regarding gender-related advantages and disadvantages in professional development. BMC Nurs. 2021;20(1):24. doi: 10.1186/s12912-021-00539-w

16. Falatah R, Salem OA. Nurse turnover in the Kingdom of Saudi Arabia: An integrative review. J Nurs Manag. 2018;26(6):630–638. doi: 10.1111/jonm.12603

17. Lazarus R, Folkman S. Stress, appraisal, and coping. New York, USA: Springer; 1984.

18. Jung HY, Lee HJ. Work experiences of nurses working as 119 paramedics. Korean J. Occup. Health Nurs. 2010, 19, 128–139 [cited 2022 Sep 20]. Available from: https://www.earticle.net/Article/A131988

19. Lee JH, Park JS. Nurses’ experience in nursing practice in correctional facilities. Qual Res 2012;13(2):92–104. doi: 10.22284/qr.2012.13.2.92.

20. Kwon JO, Oh J, Kim EH, Hahn DD. Professional identity of elementary school health teachers: A grounded theory approach. Child Health Nurs Res. 2015;21(1):64–73. doi: 10.4094/CHNR.2015.21.1.64

21. Corbin J, Strauss AL. Basics of qualitative research: Techniques and procedures for developing grounded theory. 3rd ed. Thousand Oaks, CA: Sage; 2008. doi: 10.4135/9781452230153

22. Torabi M, Borhani F, Abbaszadeh A, Atashzadeh-Shoorideh F. Experiences of pre-hospital emergency medical personnel in ethical decision-making: A qualitative study. BMC Med Ethics. 2018;19(1):95. doi: 10.1186/s12910-018-0334-x

23. Guba EG, Lincoln YS. Effective evaluation: Improving the usefulness of evaluation results through responsive and naturalistic approaches. San Francisco, CA: Jossey-Bass Publishers; 1981.

24. Kim H, Lee J. [Turnover Experience of Male Nurses]. J Korean Acad Nurs. 2017;47(1):25–38. doi: 10.4040/jkan.2017.47.1.25

25. Hwang ES. Chosun Media. Mr. Nightingale! The era of twenty thousand soon. Chosun [cited 2022 Sep 20]. Available from: http://news.chosun.com/site/data/html_dir/2014/04/24/2014042402440.html

26. Chegini Z, Asghari Jafarabadi M, Kakemam E. Occupational stress, quality of working life and turnover intention amongst nurses. Nurs Crit Care. 2019;24(5):283–289. doi: 10.1111/nicc.12419

27. Kim SO, Kang YA prediction model on the male nurses’ turnover intention. Korean J Adult Nurs. 2016;28(5):585–594. doi: 10.7475/kjan.2016.28.5.585

28. Lee KJ, Kim M. The relationship of gender role conflict and job satisfaction upon organizational commitment in male nurses. Korean J Adult Nurs. 2014;26(1):46–57. doi: 10.7475/kjan.2014.26.1.46

29. Vatandost S, Oshvandi K, Ahmadi F, Cheraghi F. The challenges of male nurses in the care of female patients in Iran. Int Nurs Rev. 2020;67(2):199–207. doi: 10.1111/inr.12582

30. Kang JH. Factors affecting social support, emotional exhaustion and job stress on job satisfaction and intention to leave of male nurses. J Korean Acad Soc Home Health Care Nurs. 2018;25(2):75–183. doi: 10.22705/jkashcn.2018.25.2.175

31. Yun JY, Ham OK, Cho IS, Lim JY. Effects of health promoting behaviors and mental health status of shift and non-shift nurses on quality of life. J Korean Public Health Nurs. 2012;26(2):268–279. doi: 10.5932/JKPHN.2012.26.2.268

32. Kang HL, Kim JH. 119 Paramedic internship experience of fourth-year nursing students. J Empl Career. 2016;6(1):1–22 [cited 2022 Sep 20]. Available from: https://www.earticle.net/Article/A270478

33. Finn P. Addressing correctional officer stress: Programs and strategies. Washington, DC, USA: National Institute of Justice; 2000.

34. Li N, Zhang L, Xiao G, Chen J, Lu Q. The relationship between workplace violence, job satisfaction and turnover intention in emergency nurses. Int Emerg Nurs. 2019;45:50–55. doi: 10.1016/j.ienj.2019.02.001

35. Altuntaş S, Harmanci Seren AK, Alaçam B, Baykal Ü. The relationship between nurses’ personality traits and their perceptions of management by values, organizational justice, and turnover intention. Perspect Psychiatr Care. 2022;58(3):910–918. doi: 10.1111/ppc.12873

36. Molavynejad S, Babazadeh M, Bereihi F, Cheraghian B. Relationship between personality traits and burnout in oncology nurses. J Family Med Prim Care. 2019;8(9):2898–2902. doi: 10.4103/jfmpc.jfmpc_423_19

37. Mariani R, Renzi A, Di Trani M, Trabucchi G, Danskin K, Tambelli R. The impact of coping strategies and perceived family support on depressive and anxious symptomatology during the coronavirus pandemic (COVID-19) lockdown. Front Psychiatry. 2020;11:587724. doi: 10.3389/fpsyt.2020.587724

38. Şahin DS, Özer Ö, Yanardağ MZ. Perceived social support, quality of life and satisfaction with life in elderly people. Educ Gerontol. 2019;45(1):69–77. doi: 10.1080/03601277.2019.1585065

39. Chen J, Li J, Cao B, Wang F, Luo L, Xu, J. Mediating effects of self□efficacy, coping, burnout, and social support between job stress and mental health among young Chinese nurses. J Adv Nurs. 2020;76(1):163–173. doi: 10.1111/jan.14208

40. Orgambídez-Ramos A, de Almeida H. Work engagement, social support, and job satisfaction in Portuguese nursing staff: A winning combination. Appl Nurs Res. 2017;36:37–41. doi: 10.1016/j.apnr.2017.05.012

41. Bajwa NM, Bochatay N, Muller-Juge V, Cullati S, Blondon KS, Junod Perron N, Maître F, Chopard P, Vu NV, Kim S, Savoldelli GL, Hudelson P, Nendaz MR. Intra versus interprofessional conflicts: Implications for conflict management training. J Interprof Care. 2020;34(2):259–268. doi: 10.1080/13561820.2019.1639645

42. Woo CH, Park JY, Kim NY. Factors influencing field adaptation in newly graduated nurses. J Korean Acad Psychiatr Ment Health Nurs. 2016;25(3):187–194. doi: 10.12934/jkpmhn.2016.25.3.187.

43. Dawis RV, Lofquist LH. A psychological theory of work adjustment. Minneapolis, USA: University of Minnesota Press; 1984.

44. Smith BW, Rojo J, Everett B, Montayre J, Sierra J, Salamonson Y. Professional success of men in the nursing workforce: An integrative review. J Nurs Manag. 2021;29(8):2470– 2488. doi: 10.1111/jonm.13445

45. Younas A, Ali N, Sundus A, Sommer J. Approaches of male nurses for degendering nursing and becoming visible: A metasynthesis. J Clin Nurs, 2022;31(5–6):467–482. doi: 10.1111/jocn.15958

46. Ministry of Justice. 2022 Korea Correctional Service Statistics [cited 2022 Sep 20]. Available from: http://www.moj.go.kr/moj/213/subview.do

47. Clark, M.J. Community health nursing advocacy for population health. 5th ed. New Jersey, USA: Pearson Prentice Hall; 2008.

48. Garcia CM, Schaffer MA, Schoon PM. Population-based public health clinical manual. 2nd ed. Indianapolis, USA: Sigma Theta Tau International; 2014.

