## Supplementary material for "Male nurses’ adaptation experiences after turnover to community institutions in Korea": IRB Certificate of Approval

Chonnam National University Hospital Institutional Review Board
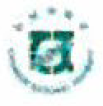

Certificate of Approval of Thesis or Dissertation

| **Recipient** | Principal Investigator | Name | Jasook Kim | College | Nursing | Degree | Ph. D. |
| --- | --- | --- | --- | --- | --- | --- | --- |
|  | Requester | Jasook Kim | | | | | |
| **IRB No.** | 1040198-180720-HR-070-03 | | | | | | |
| **Name of Research Topics** | Korean | 남자 간호사의 직종 변경 이직 과정 | | | | | |
|  | English | The Process of Job Change and Career Transition for Male Nurses | | | | | |
| **Protocol No.** |  | | | | | | |
| **Classification under the Bioethics And Safety Act** | ■ Human Subject Research □ Human Materials Research □ Genetics Research | | | | | | |
| **Types of Research** | □ Research using stored samples □ Tissue and blood research □ Research using records  □ Survey research □ Observational research □ Experimental research  ■ Other research (qualitative research using interviews) | | | | | | |
| **Date of Initial Research Proposal Approval** | March 6, 2019 | | Research Approval Period | | From the date of the initial research proposal approval  to February 28, 2020 | | |
| **Regular Reporting Interval** | □ 3 months □ 6 months □ 1 year ■ Other (none)  *The regular reporting interval cannot exceed 1 year. | | | | | | |
| **Types of Review** | ■ Expedite Review  □ Regular Review | | Date of Review | | March 6, 2019 | | |
| **Subjects of Review** | Research proposal, Description, Informed consent form | | | | | | |
| **List of Submitted Documents (Versions)** | Comparison chart of research plan revisions, version 1.2, Request for research plan review, version 1.3, Research proposal, version 1.3, Summary of the research proposal, version 1.3, Description, Informed consent form, version 1.3, Recruitment letter for research participants | | | | | | |
| **Result of Review** | - The research is approved. | | | | | | |

※ This Institutional Review Board (IRB) complies with KGCP and ICH guidelines and adheres to relevant laws such as the Bioethics And Safety Act.

※ The contents of this notification confirm that they match the records of the Institutional Review Board of Chonnam National University.

※ If there were any IRB members with conflicts of interest related to this study, they were excluded.

March 6, 2019

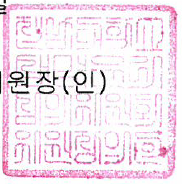

Institutional Review Board of Chonnam National University	Chairperson (Seal)
