## Supplementary material for "Male nurses’ adaptation experiences after turnover to community institutions in Korea": Clinical Studies Checklist

***PLOS ONE* Clinical Studies Checklist**

***PLOS ONE* manuscript number: PONE-D-23-10861**

| **Complete the following if your study involved human participants or human subjects’ data. These questions should be addressed for prospective and retrospective studies.** | |
| --- | --- |
| 1. | Did you obtain ethics approval for this study? - If yes, please upload (file type “Other”) the original approval document you received from your ethics committee. If the original document is in another language, please also provide an English translation. ✓ Uploaded ___ N/A - If you did not obtain ethical approval, please explain why this was not required. |
| 2. | If your study involved human participants, please report in the Methods section when participants were recruited to the study.  ✓ Completed ___ N/A |
| 3. | If you are reporting a study of medical records or archived samples, please report in the Methods section the date range in which human subjects’ data/samples were collected and the date(s) when you conducted this study.  ___ Completed ✓ N/A |
| 4. | Please specify in the Methods section whether authors had access to information that could identify individual participants during or after data collection.  ___ Completed ✓ N/A |
| 5. | If you are reporting an observational study – i.e. cohort, case-control, and cross-sectional studies – we recommend that the work is reported as per the requirements of the STROBE guidelines, and that you provide a completed STROBE checklist as a Supporting Information file with your submission.  The STROBE checklist was developed to improve the reporting of observational human subjects research, and is available here: <http://strobe-statement.org/fileadmin/Strobe/uploads/checklists/STROBE_checklist_v4_combined_PlosMedicine.docx>.  ___ Completed ✓ N/A |
| 6. | Please ensure that the author list and Corresponding Author entered in Editorial Manager match the author list and Corresponding Author in your manuscript file.  *✓* Completed |
